# Simulating the Radiological Appearance of Focal Cortical Dysplasia in MRI

**DOI:** 10.64898/2026.10.01.26364470

**Authors:** Petros Koutsouvelis, Sina Amirrajab, Leroy Volmer, Daniëlle B.P. Eekers, Olaf E.M.G. Schijns, Ralph Brecheisen, Andre Dekker

## Abstract

Focal cortical dysplasia (FCD) is among the leading causes of drug-resistant focal epilepsy and can be challenging to detect on routine MRI due to the subtle and heterogeneous appearance of lesions. Motivated by the scarcity of clinically representative cohorts, we investigate the feasibility of simulating FCD-like lesions in healthy MRI using a composition of hand-crafted image transformations. We develop *SynthFCD*, a phenomenological FCD simulator, and create 2,164 synthetic examples from healthy controls in the heterogeneous *FOMO300k* dataset. We then pretrain deep learning (DL) segmentation models on synthetic lesions and fine-tune them on real FCD cases from the University Hospital Bonn (UHB) cohort. Comprehensive evaluations show consistent improvements across simulator presets, with detection-rate gains of up to 18.5 percentage points over training from scratch and 7.4 points over self-supervised pretraining, while achieving state-of-the-art segmentation performance (0.366 DSC) on the challenging held-out UHB benchmark. These findings suggest that simulation-based synthetic data generation can bridge existing radiological knowledge and advances in DL to provide meaningful supervisory signals that could ultimately aid the detection of underrepresented and diagnostically challenging cases.

## 1 Introduction

Focal cortical dysplasia (FCD) is a congenital malformation of the cerebral cortex and one of the leading causes of drug-resistant focal epilepsy in children and adults [10]. Effective treatment typically requires complete resection or disconnection of the underlying cortical lesion, making accurate FCD localization central to successful epilepsy management [27].

Histopathologically, FCDs exhibit localized disruptions in cortical layering and are classified into Type I and Type II according to the absence or presence of atypical cells [12]. On structural MRI, a typical finding is “blurring” of the grey-to-white matter (GM–WM) boundary. Type II lesions additionally show increased cortical thickness, abnormal gyration, and cortical or subcortical signal alterations, usually T2-weighted (T2w)/fluid-attenuated inversion recovery (FLAIR) hyperintensity and mild T1-weighted (T1w) hypointensity [23]. These signal changes may extend toward the ventricles in a funnel-shaped pattern, known as the transmantle sign (TMS). Gyration-related abnormalities appear heterogeneous, ranging from enlarged adjacent cerebrospinal fluid (CSF) spaces and abnormally deep sulci to irregular cortical convolutions [6]. Type I lesions are generally more subtle but may also involve mild subcortical T2w/FLAIR hyperintensity, lobar hypoplasia, and WM atrophy [23,26].

Despite their characteristic imaging signature, FCDs are often overlooked in MRI screening due to their subtle appearance [8]. Progress in AI-assisted FCD detection aims to improve lesion “visibility” either through quantitative maps of the aforementioned radiological features [1,7,20] or by learning novel patterns directly from raw MRI using deep learning (DL) [9,17]. Though able to identify previously undetected FCDs, the scarcity of clinically representative training cohorts and the inherent variability in lesion morphology, subject anatomy, and acquisition settings compromise the transferability of AI methods to unseen populations [17]. A recent study attempted to mitigate this by synthesizing FCD lesions on healthy MRI using generative AI [14]. Despite relying on a small cohort of real FCDs, the generator produced visually plausible lesions and improved sensitivity in reduced-data settings, albeit at the cost of reduced precision.

Leveraging existing knowledge of the radiological presentation of FCDs, we propose simulating FCD-like lesions on healthy MRI via a sequence of handcrafted deformation and intensity transformations, each responsible for a distinct imaging feature. Unlike generative AI, this approach offers direct control over data generation using human-interpretable simulation parameters while eliminating reliance on real patient cohorts. This can unlock new sources of variability and enable simulation directly across scanner and site domains. We create synthetic lesions on healthy subjects from the heterogeneous *FOMO300k* dataset [4] to pretrain a DL segmentation model, which is then fine-tuned and evaluated on real FCD cases from the University Hospital Bonn (UHB) cohort [21].

Our contributions are summarized as: (i) a formulation of FCD appearance as a composition of interpretable, parametrizable transformations; (ii) *SynthFCD*, an open-source simulation platform for generating multimodal FCD-like lesions; and (iii) a downstream application achieving state-of-the-art segmentation performance on the UHB cohort and consistently outperforming self-supervised pretraining and training-from-scratch baselines.^7^

## 2 A Phenomenological Model of Focal Cortical Dysplasia

We model FCD-like lesions as an ordered composition of anatomically constrained, hand-crafted transformations that are applied to a target region of a healthy structural brain MR volume, so that the transformed image exhibits the hallmark radiological features of FCDs. The remainder of the section details the target generation process, transformation design, and the rationale driving certain modeling choices.

### 2.1 Growing the Lesion Mask

The process starts with sampling a random seed voxel along the GM–WM boundary within a given cortical region to initialize the lesion mask, optionally constrained at the bottom of a sulcus. Neighboring GM or WM voxels are added to respective priority queues *Q*_*GM*_ and *Q*_*W M*_, sorted by their physical distance *d* from the seed point and restricted to a single hemisphere. At each iteration, a voxel is popped from one of the queues and the process is repeated until a target physical volume *V* is reached. A parameter *p*_*GM*_ controls which of *Q*_*GM*_ or *Q*_*W M*_ is selected at a given iteration, while a noise factor *λ* modulates *d* prior to updating the queue. The cortical region, target volume, *p*_*GM*_ and *λ* are userprovided and control the location, size, shape, and irregularity of the resulting lesion mask. Figure 1 illustrates example masks with different parametrizations.

**Fig. 1.**
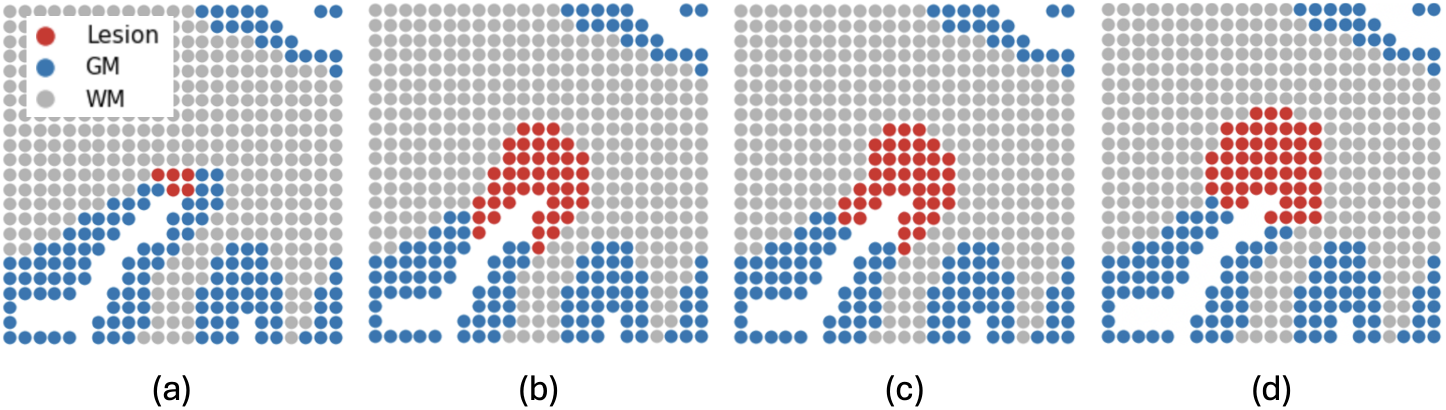
Lesion masks grown from the same seed point under different parametrizations: (a) *V* = 30 *mm*^3^, *p*_*GM*_ = 0.8, *λ* = 0.01; (b) *V* = 300 *mm*^3^, *p*_*GM*_ = 0.8, *λ* = 0.01; (c) *V* = 300 *mm*^3^, *p*_*GM*_ = 0.8, *λ* = 0.3; (d) *V* = 300 *mm*^3^, *p*_*GM*_ = 0.4, *λ* = 0.3.

### 2.2 Cortical Thickening

We model cortical thickening as the expansion of the cortex toward the WM and perpendicularly to the GM–WM interface, via the inverse displacement field

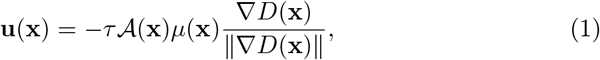

where *τ* denotes the maximum displacement in physical units, an application field decaying away from the lesion mask with rate *γ, µ* an anatomical constraint field around the inner cortical surface, *D* a signed distance field from the GM– WM boundary, and **x** the spatial coordinates of a given voxel.

The transformed image is obtained by backward warping after applying a small Gaussian smoothing kernel (*σ* ≤ 1 mm) on **u**. Parameters *τ* and *γ* are user-specified and the process may be repeated *N* times to obtain larger thickening effects while preserving smooth local deformations. The same approach applied on the pial surface can yield sulcal widening, which can emulate the lobar hypoplasia and atrophy observed in Type I lesions.

### 2.3 Grey-White Matter Boundary Blurring

GM–WM blurring is modeled as the diffusion of image intensities ℐ across the cortical boundary, governed by the equation

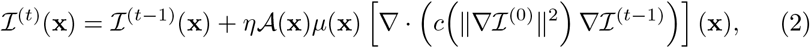

where 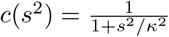 is a spatially varying Perona–Malik conduction coefficient [19] that controls sensitivity to edges and naturally provides anatomy-dependent variability. We set *κ* to the 95^*th*^ percentile of ∥∇ℐ^(0)^∥ to permit diffusion across the GM–WM junction, while retaining consistent behavior across MR contrasts. *A µ* and **x** are defined as in cortical thickening, while *η* = 0.1 denotes the update rate and *t ∈* {1, …, *N*} the current time step. We compute *c* only at *t* = 0 to ensure predictable behavior as *N* increases, with *N* controlling the strength of the blurring. The process is illustrated in Figure 2.

**Fig. 2.**
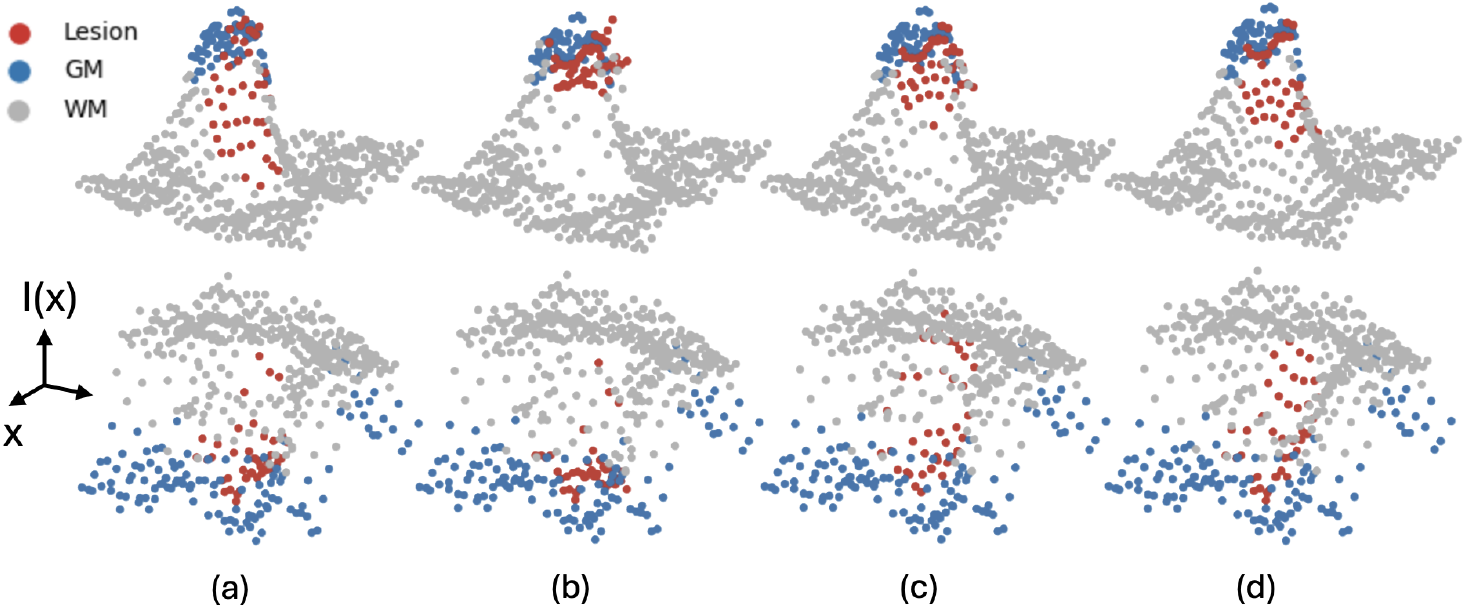
FLAIR (top) and T1w (bottom) intensities on a 2D spatial grid during cortical thickening and boundary blurring: (a) original image; (b) thickening of *τ* = 2 mm shifts intensities near the lesion toward the GM levels; (c-d) diffusion for *N* = 10 and *N* = 50 steps progressively smooths intensity gradients across the GM–WM boundary.

Repeated application of Eq. 2 can induce local texture loss. We compensate this by adding a correlated noise field *n* in affected areas, whose variance is matched to the high-frequency residuals of the original image.

### 2.4 Cortical and Subcortical Signal Alterations

An additive hyperintensity field ℋ is obtained by repeatedly diffusing a hyperintensity seed mask ℋ ^(0)^ using Eq. 2, where ℐ is replaced by ℋ, except in the computation of *c*. The initial mask ℋ ^(0)^ is generated by binarizing a correlated noise field within the WM using a spatially varying threshold that decays away from the GM–WM boundary. During diffusion, *µ* decays within the GM and is set to zero outside GM/WM regions. After N diffusion steps, ℋ is scaled to a fraction of the median lesional WM intensity and added to the original ℐ. The scaling factor, *s*, controls the extent of hyperintensity, while *s <* 0 yields hypointensity that is typically observed in T1w MRI lesions. Modeling the TMS involves estimating plausible neuronal migration trajectories from the ventricles to the cortical lesion, which is not included in this initial formulation.

### 2.5 Abnormal Gyration

Given the heterogeneity of gyration-related abnormalities, we model this phenomenon as a coordinated stochastic deformation of the cortical ribbon. Eq. 1 is first used to compute a deformation **u**, where the normal direction is replaced by correlated noise fields *n*_*x*_, *n*_*y*_, *n*_*z*_. Due to the free-form nature of the deformation, **u** is exponentiated via repeated scaling and squaring to approximate a diffeomorphic mapping and avoid artificial tearing of tissue boundaries. The number of iterations *N*, amplitude *τ*, and noise sigma *σ*_*corr*_ control the strength and spatial frequency of the resulting folding patterns.

### 2.6 Cross-Modal Coupling

Cortical deformations are shared across all modalities of a given FCD target to ensure anatomical consistency. Likewise, GM–WM blurring parameters are kept fixed. Texture restoration is independently seeded per scan to account for acquisition-specific noise and texture variations. GM/WM signal alterations share the same ℋ across modalities, while *s* remains modality-dependent: T2-like scans (e.g., T2w and FLAIR) receive a *s >* 0, which is stochastically sign-inverted and downscaled for T1w to reflect the non-trivial and under-specified T1w–T2w relationship when relying solely on radiological observations [16].

## 3 Simulation Workflow

Each run takes as input a healthy subject with spatially aligned T1w/T2-like MR volumes and an anatomical segmentation mask. Simulation parameters are sampled from predefined distributions, referred to as *presets*; physical quantities (e.g., volume *V*, thickening *τ*) are drawn from truncated lognormal distributions. Presets are chosen based on prior literature [2,5,24], intended behavior, and visual plausibility checks. We define four preset families: *universal*, covering Type I- and Type II-like FCDs; *type-II*, restricted to Type II-like FCDs; *minimal*, disabling abnormal gyration; and *extreme*, using deformation and hyperintensity parameter ranges that may exceed visual realism. Type I-like lesions are modeled as smaller lesions (*V* ≤ 1000 mm^3^), primarily in the temporal lobe, with sulcal widening instead of cortical thickening and a milder hyperintensity. The phenomenological model is then executed for each scan to generate transformed modality-aware FCD-like volumes and a corresponding synthetic lesion mask.

## 4 Experimental Setup

### 4.1 Synthetic FCD Data Generation

We searched the *FOMO300k* dataset [4] for healthy T1w volumes with at least one FLAIR from the same session. Volumes with voxel spacing exceeding 5 mm in any dimension were excluded. This yielded 2,164 T1w images from 1,370 unique subjects across 15 sources, each randomly paired with a same-session FLAIR to form 2,164 target synthetic subjects. Images were then skull-stripped with *HD-BET* [13] and preprocessed via a custom *ANTs*-based [22] pipeline, comprising bias field correction, T1w–FLAIR co-registration, affine registration to MNI space, cropping to brain size, intensity clipping, and z-score normalization.^8^ Anatomical segmentation masks with cortical parcellation were generated from the preprocessed T1w using *SynthSeg* [3]. Resulting pairs were then processed by the *SynthFCD* simulator under all four presets and two random seeds, yielding eight datasets of 2,164 synthetic FCD subjects each.

### 4.2 FCD Lesion Segmentation

Real FCD cases were collected from the UHB cohort of Type II FCDs [21], yielding 82 subjects after applying the same curation and preprocessing steps as in Section 4.1. A difficult-to-detect subset previously benchmarked against human experts [25] and DL methods [15] was held out for testing.^9^

We pretrained one segmentation model per synthetic dataset using a hybrid *Swin* encoder with stage-wise convolutions [11] and a lightweight feature-pyramid decoder [18]. Inputs were cropped to 128^3^-voxel patches. Models were optimized for 200 epochs with Dice–cross-entropy loss and AdamW (LR = 5×10^*−*4^). The network was fine-tuned on real FCDs for an additional 400 epochs, transferring all parameters. The encoder was frozen for the first 5% of updates and then optimized at 10% of the base LR. No hyperparameter tuning was performed. Baseline comparisons involved training directly on the real cohort without pretraining (*from-scratch*) and self-supervised pretraining via masked image modeling (*MIM*) [28] on the same 2,164 healthy inputs. For *MIM*, we employed a custom intra- and inter-modal masking strategy for T1w–FLAIR fusion and repeated pretraining for two seeds. Only the encoder was transferred, hence we further evaluated *SynthFCD* pretraining using a newly initialized decoder for direct comparison. All fine-tuning and *from-scratch* runs were repeated for three seeds. This yielded six final models for each *SynthFCD* preset and decoder-transfer setting, six for *MIM*, and three for the *from-scratch* baseline.^8^

**Table 1.** Mean performance across all experiments on the UHB test set.

|  | Baselines |  | *SynthFCD* (encoder-only) |  |  |  | *SynthFCD* (full transfer) |  |  |  |
| --- | --- | --- | --- | --- | --- | --- | --- | --- | --- | --- |
|  | *scratch* | *MIM* | *univ.* | *type-II* | *mini.* | *extr.* | *univ.* | *type-II* | *mini.* | *extr.* |
| DSC (%) | 23.59 | 29.90 | 32.09 | 35.84 | 32.46 | 33.61 | 31.89 | **36.57** | 34.59 | 31.78 |
| D-OVO (%) | 55.56 | 62.96 | 66.67 | 72.22 | 69.14 | 69.75 | 67.90 | 70.37 | **72.84** | 65.43 |
| D-0.22 (%) | 41.98 | 53.09 | 55.56 | 58.64 | 54.94 | 56.17 | 55.56 | **60.49** | 59.26 | 55.56 |

Segmentations were obtained via sliding-window inference over whole-brain inputs and constrained to a single output cluster. Performance was evaluated using the Dice Similarity Coefficient (DSC) and bounding-box detection rate at the one-voxel overlap (D-OVO) and 0.22 DSC (D-0.22) thresholds, as defined in [15]. Subject-wise metrics were averaged across seeds, followed by mean aggregation and 95% confidence interval (CI) estimation via bootstrapping (N=10,000).

## 5 Results and Discussion

Figure 3 shows representative synthetic data across different parametrizations, subjects, and imaging settings. Mean processing time was 4.4 seconds per simulation input on a standard CPU. Quantitative results from all experiments are reported in Table 5. *SynthFCD* pretraining consistently improved FCD segmentation and detection over the *from-scratch* and *MIM* baselines across all presets, transfer settings, and evaluation measures.

**Fig. 3.**
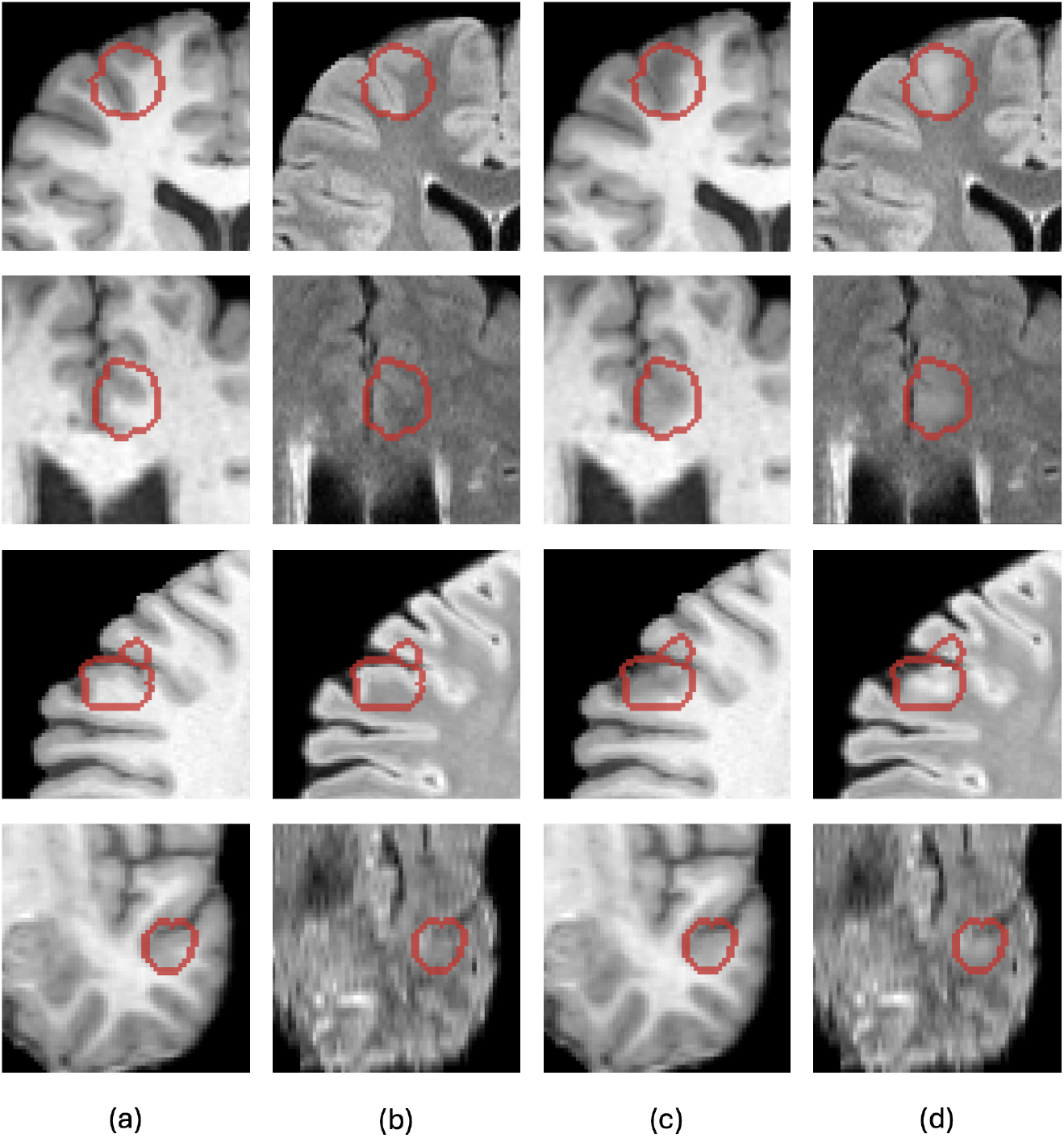
*SynthFCD* input–output pairs across subjects and imaging settings: healthy T1w/FLAIR inputs (a,b), synthetic outputs (c,d), and lesion masks outlined in red. Each row corresponds to a different subject. From top to bottom: Type II-like lesions showing pronounced cortical thickening with T1w hypointensity/FLAIR hyperintensity; cortical thickening with GM–WM blurring; enlarged adjacent CSF space with cortical/subcortical signal alterations; and a subtle Type I-like lesion with mild GM– WM blurring, increased FLAIR signal, and adjacent sulcal widening.

The *type-II* preset with full-network transfer achieved superior overall performance, with a DSC of 0.366 (95% CI: 0.259–0.474) and D-0.22 of 0.605 (0.432–0.778). This is a substantial improvement over training from scratch, with a DSC of 0.236 (0.146–0.332) and D-0.22 of 0.420 (0.272–0.568), and selfsupervised pretraining, with a DSC of 0.299 (0.198–0.407) and D-0.22 of 0.531 (0.360–0.704). Notably, all models pretrained with *SynthFCD* data exceeded the previously reported state-of-the-art DSC of 0.30 on this benchmark^10^, which included *MAP18, MELD*, and other established methods [15], despite fine-tuning on only a small real FCD cohort. All pretrained models, including *MIM*, exceeded the reported human-expert D-0.22 of 0.49 [25]. Interestingly, synthetic pretraining alone identified some real FCDs without fine-tuning, but with limited utility and no clear preference across presets (DSC: 0.037–0.047; D-OVO: 0.092–0.220).

The findings suggest that phenomenologically simulated FCD lesions could provide meaningful supervision for identifying real FCDs beyond that of generalist methods under the same data budget. However, more experiments are needed to establish how these benefits scale with increasing pretraining size or the representativeness of real FCD cohorts. Results further imply that transformation design and simulation parametrization can also play an important role, highlighted by the improved performance upon dropping the more under-specified Type I-like lesions (*type-II* vs. *universal*) and stochastic cortical deformations (*minimal* vs. *universal*). Whether such fluctuations can be attributed to synthetic data–real cohort alignment (e.g., the superior performance of *type-II* on the Type II UHB cohort) or other confounding factors (e.g., subtler lesions providing ambiguous supervision signals) remains to be seen in larger and more diverse real patient cohorts. Lastly, the present simulation workflow does not incorporate all radiological cues (e.g., TMS) or parameter interactions (e.g., lesion location and cortical thickening). The extent to which visual realism and mechanistic plausibility in the generation process strengthen the downstream utility of the simulated datasets is an open question and should be further explored.

## 6 Conclusion

We introduced *SynthFCD*, a phenomenological simulator of FCD-like lesions, and demonstrated its potential to improve the localization of real FCDs. This line of work may reveal new ways of translating radiological knowledge into effective machine supervision, ultimately aiding the detection of subtle lesions while bridging the gap between technical and clinical research.

## Data Availability

All data produced are available online at https://huggingface.co/datasets/FOMO-MRI/FOMO300K, https://openneuro.org/datasets/ds004199/versions/1.0.6, and https://github.com/MaastrichtU-CDS/SynthFCD.

## Acknowledgments

This publication is part of the AIM@EPILEPSY project (No. KICH1.ST03.21.016), funded in part by the Dutch Research Council (NWO) under the KIC–MISSION 2021 program, and is further supported by the Dutch national e-infrastructure through the SURF Cooperative (No. EINF-13271).

## Footnotes

7 Code and model weights: https://github.com/MaastrichtU-CDS/SynthFCD.

8 Implementation details and experiment configurations are provided with the code.

9 One test subject (sub-00074) did not meet the spacing criterion and was excluded.

10 At the time of writing in July, 2026. As of October 1, 2026, the claim still holds.

